# Genome-wide association studies of hospitalized influenza identify 4 risk loci and a relationship with COVID-19

**DOI:** 10.1101/2025.04.25.25325850

**Authors:** Xiao Xue, Pui-Ching Chan, Hon-Cheong So

**Affiliations:** School of Biomedical Sciences, The Chinese University of Hong Kong, Shatin, Hong Kong, China; KIZ-CUHK Joint Laboratory of Bioresources and Molecular Research of Common Diseases, Kunming Institute of Zoology and the Chinese University of Hong Kong, China; Department of Psychiatry, The Chinese University of Hong Kong, Hong Kong, China; CUHK Shenzhen Research Institute, Shenzhen, China; Margaret K.L. Cheung Research Centre for Management of Parkinsonism, The Chinese University of Hong Kong, Shatin, Hong Kong, China; Brain and Mind Institute, The Chinese University of Hong Kong, Hong Kong SAR, China; Hong Kong Branch of the Chinese Academy of Sciences Center for Excellence in Animal Evolution and Genetics, The Chinese University of Hong Kong, Hong Kong SAR, China

## Abstract

Influenza remains a significant global health burden, with the potential for severe complications and mortality. This study investigates the genetic architecture of hospitalized influenza and explores its shared and distinct genetic factors with hospitalized COVID-19. A binary GWAS of hospitalized influenza using UK Biobank data, followed by meta-analysis with FinnGen, identified three risk loci: ST6GAL1, AASDHPPT, and SPATS2L. Additionally, a time-to-event GWAS revealed a novel susceptibility locus, UGT2B4. Further differentiation analysis with hospitalized COVID-19 identified 29 differentially associated loci, highlighting their distinct genetic architectures. Colocalization analysis uncovered shared genetic mechanisms, with ST6GAL1 and ICAM5 emerging as key candidates. These findings provide new insights into the genetic basis of influenza and its relationship with COVID-19, offering potential therapeutic targets and avenues for personalized medicine.

## Introduction

Influenza is an acute respiratory infection that affects millions of people worldwide, causing 3 to 5 million severe cases and 290000 to 650000 deaths annually^1^. While often self-limiting with recovery in 2 to 7 days for healthy individuals, influenza can lead to a range of medium- and long-term complications affecting multiple organ systems, including the respiratory, cardiovascular, digestive, and neurological systems^2^. Certain populations, such as young children, pregnant women, older adults, and people with chronic conditions, are at a higher risk of hospitalization and mortality^3, 4^.

Recent studies have increasingly recognized the role of genetic factors in influenza severity and susceptibility. For example, the rs12252-C allele at the IFITM3 locus was found to be associated with increased disease risks in both Han Chinese and Caucasian populations, based on allele frequency comparison tests between infected and healthy individuals^5, 6^. Functional studies have further confirmed the protective role of IFITM3 against influenza viral infection^5, 6^. Similarly, a GWAS with 425 Chinese participants identified an association between severe influenza and the rs2564978-T variant in the CD55 gene^7^. Despite these important findings, few GWAS have explicitly targeted hospitalized influenza cases, leaving a critical gap in our knowledge of the genetic determinants behind severe disease outcomes.

To broaden our understanding, it is also valuable to consider related respiratory infections such as COVID-19, which shares similar transmission pathways and clinical symptoms with influenza^8^. However, the two diseases are caused by different viruses and have different etiological characteristics. For instance, human influenza viruses prefer α-2,6-linked sialic acid receptors for cell entry, whereas SARS-CoV-2 relies on ACE2 and TMPRSS2. Although a recent comparative analysis of self-reported influenza and COVID-19 GWAS highlighted their distinct genetic architectures^9^, the relationship between severe disease outcomes, particularly hospitalization, remains unclear.

In this study, we addressed these knowledge gaps by characterizing the genetic landscape of hospitalized influenza and exploring its shared and differentiated genetic features with hospitalized COVID-19. We conducted both binary and time-to-event GWAS analyses to identify loci associated with the risk of influenza-related hospitalization. An integrative gene prioritization framework was then applied to find potential causal genes underlying these associations. We further performed genetic differentiation and colocalization analyses to determine commonalities and distinctions between hospitalized influenza and hospitalized COVID-19. Collectively, our findings provide novel insights into the host genetic factors of hospitalized influenza and its relationship with hospitalized COVID-19, which may help to clarify their respective pathophysiology and inform targeted therapeutic approaches.

## Methods

### Genome-Wide Association Study (GWAS) Analysis

We conducted a GWAS on hospitalized influenza (Flu_UKBB) using individual-level phenotypic and genotypic data from the UK Biobank (UKBB)^10^. Cases were defined as UKBB participants with hospitalization or death records due to influenza (ICD-10 codes: J09, J10, J11; HESIN-restricted, UKBB Category 2000), while all others served as controls. To minimize population stratification bias, we included only White British individuals of Caucasian ancestry, as defined in UKBB Data-Field 22006, resulting in 1,037 cases and 404,108 controls. Genetic variants with an imputation score >0.3 were retrieved and subjected to quality control using PLINK2^11^, applying a Hardy-Weinberg equilibrium test threshold of P < 1×10⁻⁶ and a missing call rate <0.01. GWAS was conducted using fastGWA-GLMM^12^, which efficiently fits a mixed-effects logistic regression model for binary traits in large-scale biobank data and applies Saddle Point Approximation (SPA) to correct for case-control imbalance. Covariates, including age, sex, age², age × sex, and the first 10 principal components, were adjusted for in the analysis. We standardized GWAS summary statistics using MungeSumstats^13^ with default parameters, aligning them to GRCh37. After filtering, 7,074,802 bi-allelic SNPs with a minor allele count (MAC) >10 present in the reference file (dbSNP144) were retained for further meta-analysis.

### Time-to-Event GWAS

To account for family relatedness in survival analysis, we removed 48,034 third-degree relatives (gcta --grm-cutoff 0.125) from a cohort of 405,145 genetically confirmed Caucasians with phenotypic, genotypic, and clinical data updated to 2021 in the UKBB. For each unrelated participant, those with at least one influenza hospitalization record were assigned event = 1, with survival time (years) calculated as the difference between the first diagnosis date and birth date. For all others, event = 0, and survival time was determined as the minimum of death date or lost-to-follow-up date, minus birth date. The same covariates and genotype quality control criteria as in Flu_UKBB were applied. The genome-wide time-to-event analysis was conducted using SPACox^14^, an SPA-based Cox proportional hazards model, to identify SNPs associated with hospitalized influenza (Flu_SPACox).

### Meta-Analysis Using Effect Sizes

A genome-wide meta-analysis combining UKBB and FinnGen (release 11) data was performed using METAL^15^ under an inverse-variance weighted fixed-effects model. FinnGen summary statistics (Flu_FinnGen) for clinically diagnosed influenza (endpoint: J10_INFLUENZA; 10,134 cases, 378,292 controls) were downloaded. After standardizing the GWAS formats for UKBB and FinnGen, overlapping variants with MAC >10 were meta-analyzed and subsequently used as input for dist^16^ to impute unmeasured SNPs. Following quality control (MAC >10, imputation quality INFO >0.8), we obtained 8,258,810 bi-allelic SNPs, comprising 7,241,071 common variants (minor allele frequency [MAF] >0.01; Flu_meta) and 1,017,739 rare variants (MAF <0.01).

### Meta-Analysis Using P-Values

To improve statistical power while controlling the false positive rate, P-values from Flu_SPACox and Flu_meta were combined using the harmonic mean P-value (HMP) method^17^. This analysis yielded 6,212,866 SNPs with MAF >0.01 (Flu_hmp), which were meta-analyzed using the R package *harmonicmeanp*.

### Estimation of SNP-Based Heritability and Genetic Correlation

We first merged GWAS results with HapMap3 SNPs to obtain a set of stable, high-quality variants. LD score regression (LDSC)^18^ was then used to estimate SNP-based heritability (h²) on the liability scale and genetic correlation (rg) using precomputed 1000 Genomes European LD scores. The case ratio was assumed to represent both sample and population prevalence.

### Functional Mapping and Annotation of GWAS

All GWAS summary statistics were annotated and characterized using the FUMA^19^ online platform. Genomic risk loci were identified via the SNP2GENE module with default settings. Additionally, MAGMA^20^ was used for gene and gene-set analyses without extending genic boundaries to detect phenotype-associated genes and biological pathways. MAGMA gene-property analysis integrated GWAS with gene expression data from 54 tissue-specific datasets in GTEx v8 and 12 single-cell expression datasets, identifying enriched tissues and cell types. Multiple testing correction was applied using a false discovery rate (FDR) <0.05.

### Transcriptome-Wide Association Study (TWAS) and Splicing TWAS (spTWAS)

Tissue-specific TWAS and spTWAS analyses were conducted to identify genes whose genetically predicted mRNA expression or splicing activity is associated with hospitalized influenza. We first integrated GWAS data with GTEx v8 eQTL/sQTL MASHR models for 49 tissues using S-PrediXcan^21^. To enhance statistical power, we then performed a cross-tissue TWAS/spTWAS using S-MultiXcan^22^. Intron coordinates were mapped to genes using leafcutter^23^ for improved interpretability.

### Proteome-Wide Association Study (PWAS)

To detect risk genes whose genetically regulated protein abundance is associated with hospitalized influenza, we conducted a PWAS using FUSION^24^. Predictive models of plasma protein levels from European American populations^25^ were combined with GWAS data to estimate gene-phenotype associations.

### Integrative Gene Prioritization Framework

We employed an integrative framework to prioritize risk genes within genome-wide significant loci by combining multiple GWAS annotation tools and gene-based association tests. First, Open Targets^26^ was used to identify the closest gene and the gene with the highest V2G score for each significant SNP. The V2G pipeline integrates various datasets, including QTLs, chromatin interactions, physical distances, and computational functional predictions, to rank potential causal genes. FUMA^19^ was then applied to annotate influenza GWAS results. SNPs were assigned to genes based on significant eQTL mapping (FDR <0.05), with the gene showing the strongest eQTL signal (minimum FDR) designated as the most relevant. SNPs were further characterized using CADD scores^27^ (variant deleteriousness) and RegulomeDB scores^28^ (regulatory potential). Functional enrichment was assessed using ANNOVAR^29^. Genes annotated with the highest CADD scores, lowest RegulomeDB ranks, or nonsynonymous substitutions were prioritized. Additionally, significant genes from MAGMA, TWAS, spTWAS, and PWAS analyses were incorporated. Finally, PheWAS was performed using GWAS Atlas^30^ to test gene associations with immunological or respiratory traits.

### Identification of Differentially Associated SNPs and Genes

To identify genetic markers distinguishing hospitalized influenza from COVID-19, we applied the DDx algorithm^31^, treating hospitalized influenza as the pseudo-case and COVID-19 as the pseudo-control. GWAS data for hospitalized COVID-19 were obtained from the COVID-19 Host Genetics Initiative (Release 7, B2_ALL, primarily European ancestry). Effect sizes of differential associations between the two conditions were calculated using original GWAS effect sizes. Further analysis of genes, pathways, tissues, and cell types was conducted using MAGMA, TWAS, spTWAS, and PWAS.

### Identification of Colocalized SNPs and Genes

To identify genetic variants contributing to both hospitalized influenza and COVID-19 risk, we applied cofdr^32^ using GWAS Z-scores as input. The posterior probability (PPA) was computed to classify SNPs as influenza-specific, COVID-19-specific, shared (PPA >0.9), or unrelated. Cofdr was also extended to the gene level using Z-scores from MAGMA, TWAS, spTWAS, and PWAS. For further methodological details, refer to ref^32^ and the supplementary materials.

## Results

### Genome-Wide Risk Loci Associated with Hospitalized Influenza

We conducted a genome-wide association study (GWAS) on hospitalized influenza in the UK Biobank (UKBB) and subsequently performed a meta-analysis incorporating the GWAS of clinically ascertained influenza from FinnGen Release 11. Z-scores for SNPs that were not directly measured in the meta-analysis were imputed and filtered using an INFO score threshold of >0.8.

Applying FUMA’s default LD-clumping parameters, we identified eight independent significant SNPs across four genomic risk loci in Flu_meta, six of which were imputed. However, rs139529053 was the only SNP at its respective locus, and no surrounding common variants were found to be in linkage disequilibrium (LD) with it. Therefore, it was excluded as a lead SNP (Table 1; Figure 1c). Among the remaining three loci, one (rs117391169-G) increased the odds of hospitalized influenza by approximately 10.7% (BETA = 0.044), while the others were protective.

**Figure 1.**
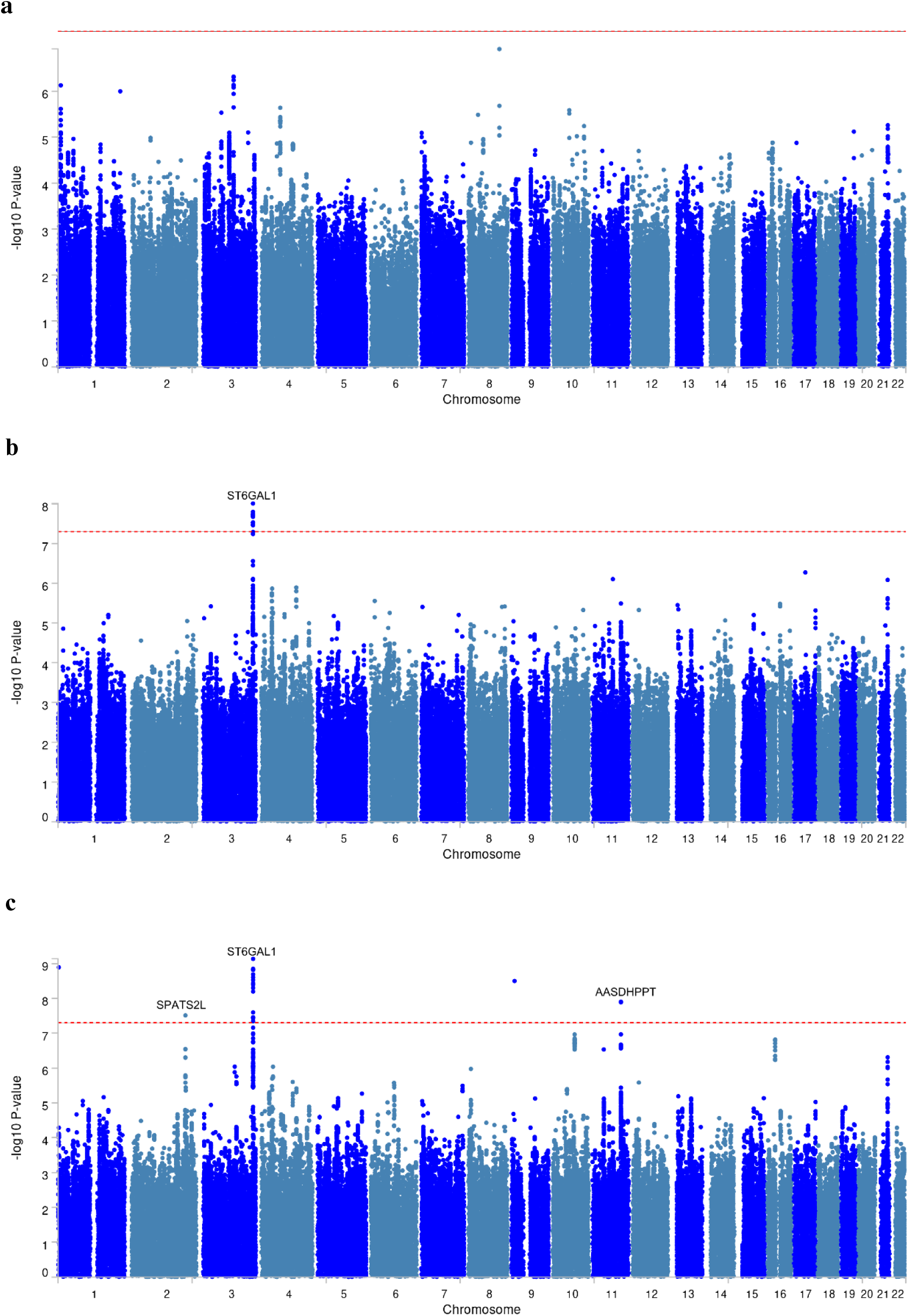
Manhattan Plot of the Common Variant GWAS for Hospitalized Influenza. Each data point in the Manhattan plot represents the −log₁₀ P-value of a single nucleotide polymorphism (SNP) with a minor allele frequency (MAF) > 0.01 from (a) UK Biobank (UKBB), (b) FinnGen, and (c) a meta-analysis of UKBB and FinnGen data. The red horizontal line indicates the conventional genome-wide significance threshold (P < 5 × 10⁻⁸). The nearest protein-coding gene to each lead SNP (within ±500 kb, as defined by Open Targets) is labeled in the plot.

**Table 1.**
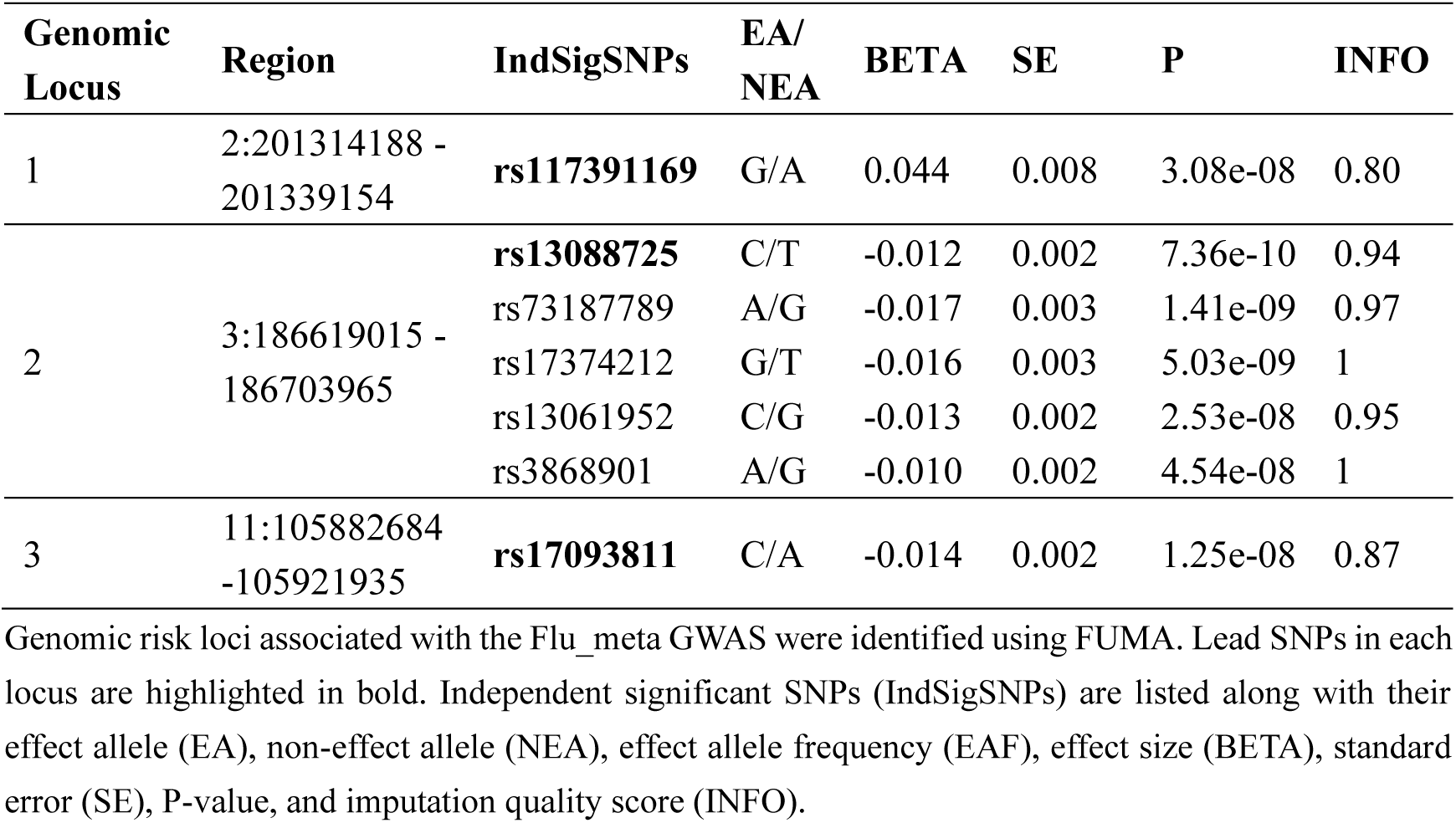
Genomic Risk Loci Associated with Hospitalized Influenza.

The strongest association was observed for rs13088725, an intronic variant of ST6GAL1, which encodes β-galactoside α-2,6-sialyltransferase I, an enzyme responsible for the synthesis of α-2,6-linked sialic acid receptors^33^. A phenome-wide association study (PheWAS) in the GWAS Atlas database^30^ indicated that the rs13088725-C allele was associated with the inflammatory marker mean platelet volume (MPV)^34^ at an FDR threshold of 0.05 (P = 6.31×10⁻⁴, FDR = 0.013). Additionally, four other independent significant SNPs were identified within this locus, including rs17374212 and rs3868901, both of which were present in the UKBB and FinnGen cohorts and passed heterogeneity tests. Specifically, rs17374212 was nominally significant in Flu_UKBB (P = 0.0173) and reached suggestive significance in Flu_FinnGen (P = 5.78×10⁻⁸), while rs3868901 showed a similar pattern (P = 0.0481 in UKBB; P = 2.77×10⁻⁷ in FinnGen). These influenza-associated SNPs also functioned as significant expression quantitative trait loci (eQTLs) for ST6GAL1 in various tissues, as identified using the GTEx^35^ and eQTLGen^36^. For instance, rs13088725-C and rs13061952-C were associated with lower ST6GAL1 expression in blood (P = 5.5×10⁻¹¹)^36^ and lung (P = 1.5×10⁻⁵)^35^, respectively.

The second most significant association was observed for rs17093811, located near AASDHPPT, which encodes the phosphopantetheinyl transferase enzyme^37^. PheWAS revealed that this SNP was correlated with a human CD4 T-cell subset, CD28⁺CD127⁻ (P = 1.96×10⁻⁴, FDR = 0.021)^38^. Additionally, the rs17093811-C allele was linked to higher AASDHPPT expression in blood (P = 7.9×10⁻²³)^36^.

Another genome-wide significant association was identified for rs117391169, located within intronic regions of SPATS2L, a type I interferon-stimulated gene implicated in multiple autoimmune diseases^39, 40^. According to PheWAS, this SNP was associated with prospective memory, a cognitive ability related to remembering planned tasks (P = 3.22×10⁻⁶, FDR = 0.00027)^41^.

### A Novel Locus Identified in the Genome-Wide Survival Analysis of Hospitalized Influenza

We applied SPACox to the hospitalized influenza cohort from the UK Biobank (UKBB) and identified three genome-wide significant SNPs, all mapping to a single locus (Flu_SPACox; Table 2; Figure 2a). After integrating P-values from Flu_SPACox and Flu_meta using the harmonic mean P-value (HMP) approach, we identified four independent SNPs across two loci (Flu_hmp; Table 3; Figure 2b). Notably, the ST6GAL1 locus was detected in both the conventional binary GWAS (Flu_meta) and the meta-analyzed time-to-event GWAS (Flu_hmp).

**Figure 2.**
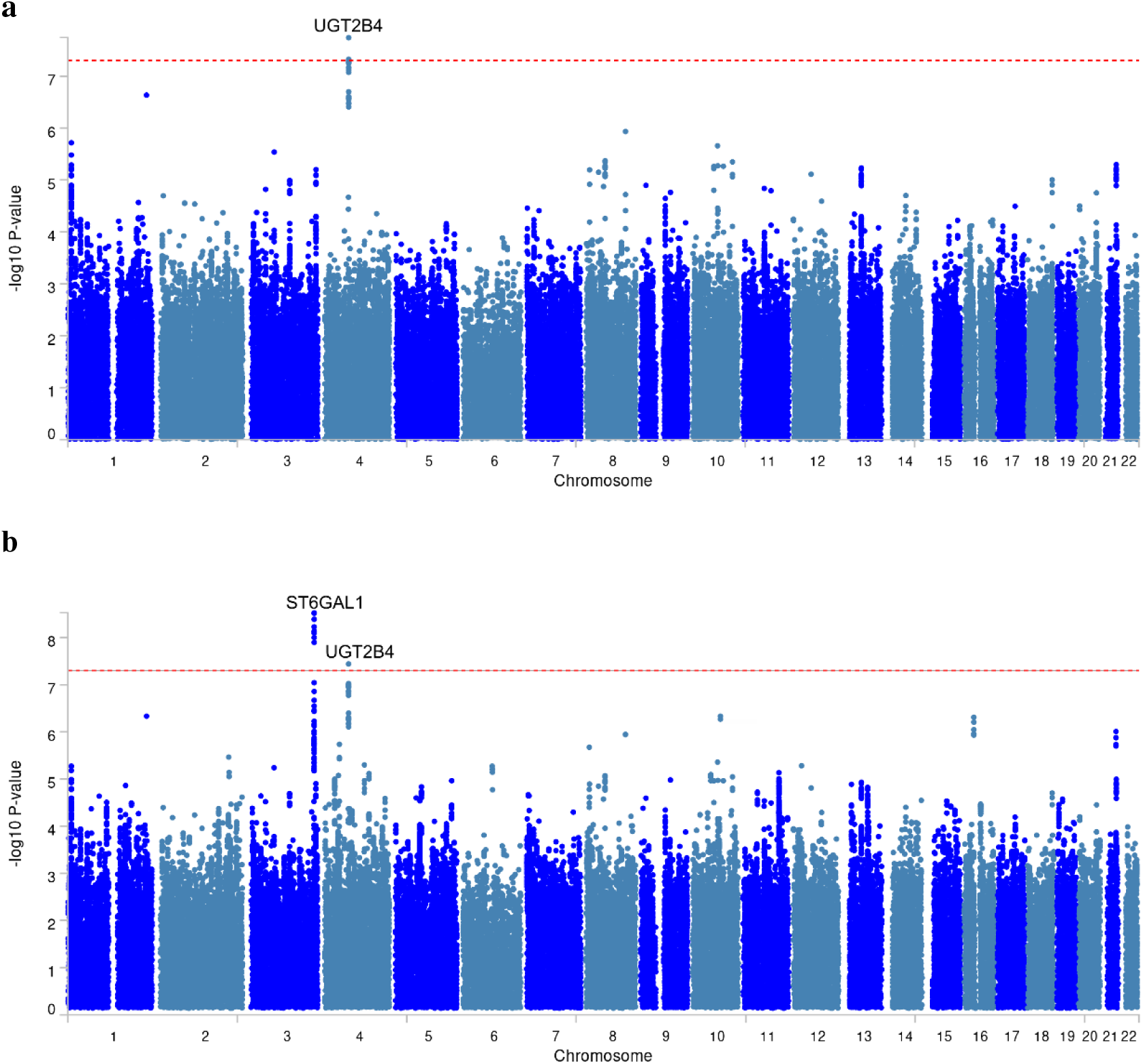
Manhattan Plot of the Time-to-Event GWAS for Hospitalized Influenza. Manhattan plots were generated using P-values calculated from (a) SPACox and (b) the harmonic mean P-value (HMP) approach.

**Table 2.**
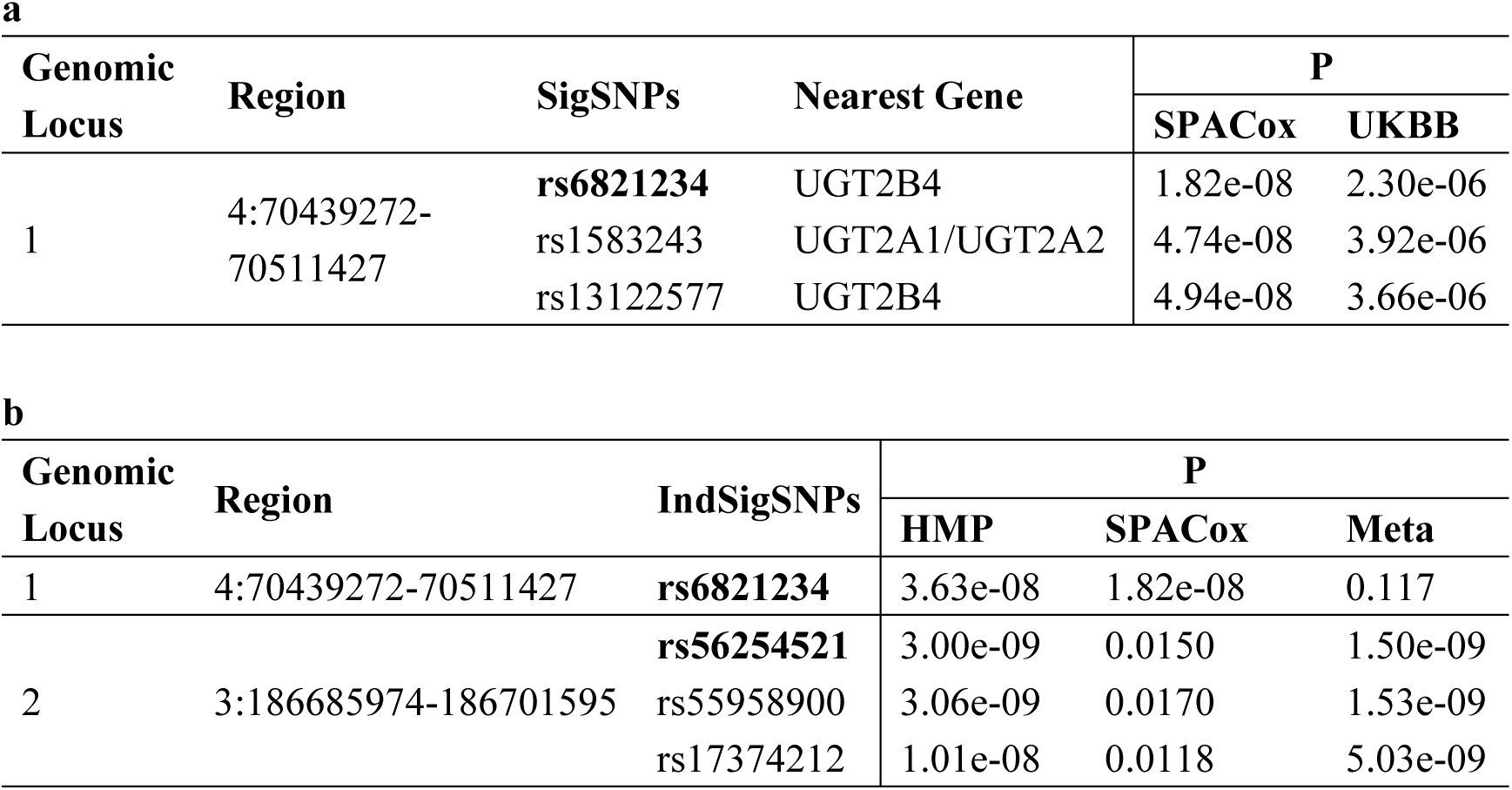
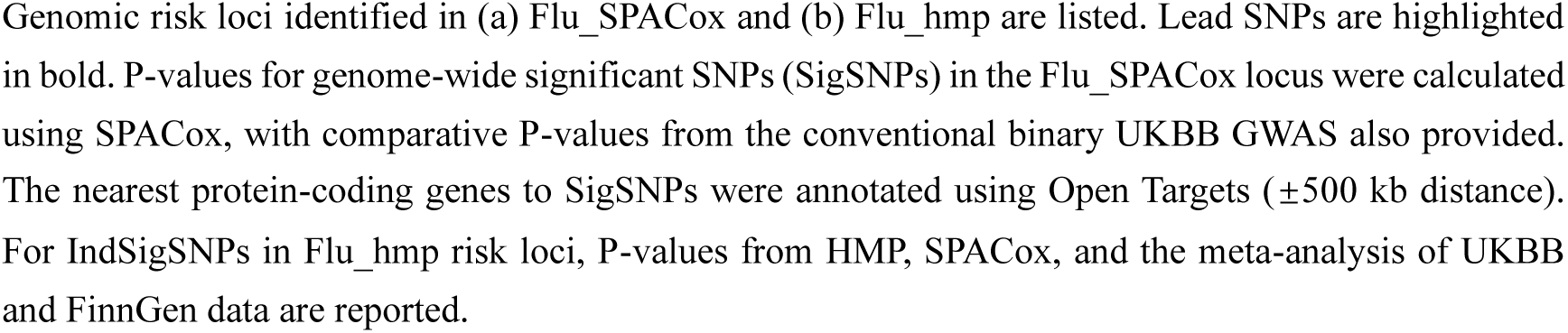
Genomic Risk Loci Associated with the Time-to-Event GWAS of Hospitalized Influenza.

**Table 3.**
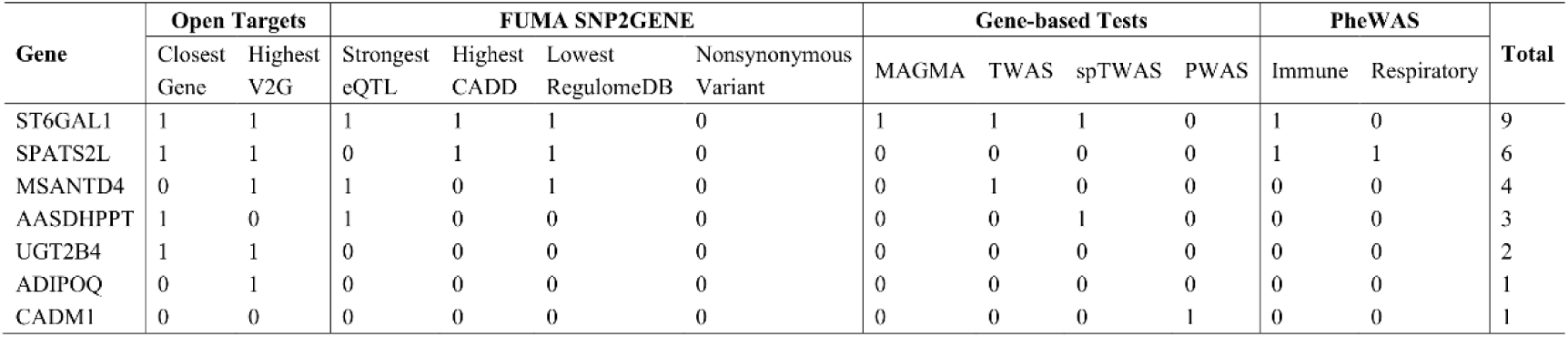
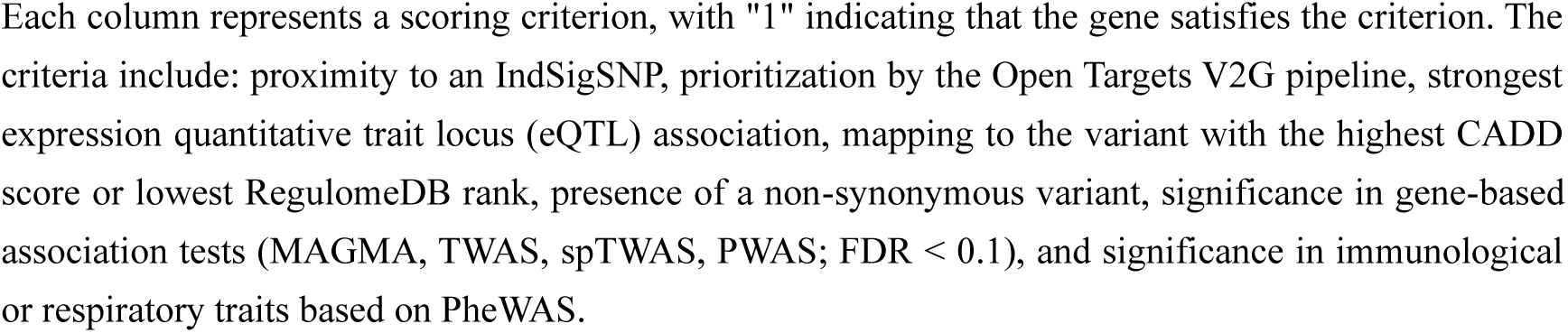
Prioritization of Risk Genes Associated with Hospitalized Influenza.

A novel locus, rs6821234, was identified through a genome-wide survival analysis of hospitalized influenza patients. This SNP is located closest to UGT2B4, with UGT2A1 and UGT2A2 being the second and third nearest genes, respectively. Rs6821234 has been shown to directly influence the expression levels of various isoforms within the UGT2A and UGT2B gene families, including UGT2A1 in the liver^35^, UGT2A3P7 in the left ventricle of the heart^35^, UGT2B4 in the lung^35^, and UGT2B11 in the blood^36^. Additionally, a phenome-wide association study (PheWAS) revealed a significant association between rs6821234 and TNF-α levels in the immune system (P = 1.95×10⁻⁴, FDR = 0.013)^42^.

### Gene Prioritization Suggests Potential Targets for Influenza Therapy

The gene prioritization framework was applied to genes that met at least one of the following criteria: (1) the closest gene to independent significant SNPs in the meta-analyzed influenza GWAS or time-to-event influenza GWAS, (2) the gene with the highest variant-to-gene (V2G) score for these SNPs, or (3) genes identified in any of the gene-based association tests.

Among the prioritized genes, ST6GAL1 received the highest prioritization score, making it the strongest candidate for association with hospitalized influenza. Transcriptome-wide association study (TWAS) analysis linked elevated ST6GAL1 mRNA expression in seven tissues to an increased risk of hospitalized influenza (Supplementary Table 3), including the thyroid, sigmoid colon, testis, left ventricle, atrial appendage, tibial artery, and minor salivary gland. Additionally, splicing TWAS (spTWAS) identified a significant association between splice activity at chr3:186681714-186756530 within ST6GAL1 and hospitalized influenza in prostate tissue (Supplementary Table 4). Furthermore, phenome-wide association study (PheWAS) analysis demonstrated significant associations between ST6GAL1 and various types of blood cells^34^.

SPATS2L ranked second in prioritization and was associated with pulmonary function^43^, pain in throat and chest^41^ and red blood cells^34^, as indicated by PheWAS. MSANTD4 was prioritized by the V2G pipeline in Open Targets, despite not being the nearest gene to the lead SNP rs17093811 on chromosome 11. TWAS identified MSANTD4 as significantly associated with gene expression in the liver, small intestine terminal ileum, and esophagus mucosa (Supplementary Table 3). Conversely, AASDHPPT was the closest gene to the lead SNP rs17093811 and had the second-highest V2G score. SpTWAS analysis in whole blood suggested its involvement in hospitalized influenza through splicing regulation (Supplementary Table 4). UGT2B4 satisfied two prioritization criteria and was the only gene identified in the time-to-event GWAS of hospitalized influenza.

Finally, ADIPOQ and CADM1 received the lowest prioritization scores. ADIPOQ had the highest V2G score for the independent SNP rs17374212 on chromosome 3, despite being the second closest gene to the SNP. CADM1 was the only gene identified by proteome-wide association study (PWAS) and exhibited protective effects against hospitalized influenza (Supplementary Table 5).

### Differentiation Analysis of Hospitalized Influenza and COVID-19 at the SNP and Gene Levels

To identify distinct genetic architectures underlying hospitalized influenza and COVID-19, we applied the DDx method to compare the GWAS results of these two diseases (Flu_meta vs. B2_ALL). Our analysis identified 3,019 SNPs that were differentially associated with the two conditions at a stringent P-value threshold of 5×10⁻⁸, aggregating into 29 risk loci (Table 4a; Figure 3). Notably, all identified lead SNPs reached genome-wide significance in COVID-19, except for rs2876034 and rs12567716, which achieved significance at P < 5×10⁻⁷. In contrast, none of these SNPs showed significant associations with influenza, even at the more lenient suggestive significance level of P < 1×10⁻⁵ (Table 4b). Furthermore, only two SNPs— rs34725611-G (BETA = −0.084, P = 0.023) and rs10774679-T (BETA = −0.076, P = 0.011)— marginally reached nominal significance (P < 0.05) in the GWAS for hospitalized influenza. Both SNPs conferred protective effects against hospitalized influenza but exhibited opposite effects in hospitalized COVID-19.

**Figure 3.**
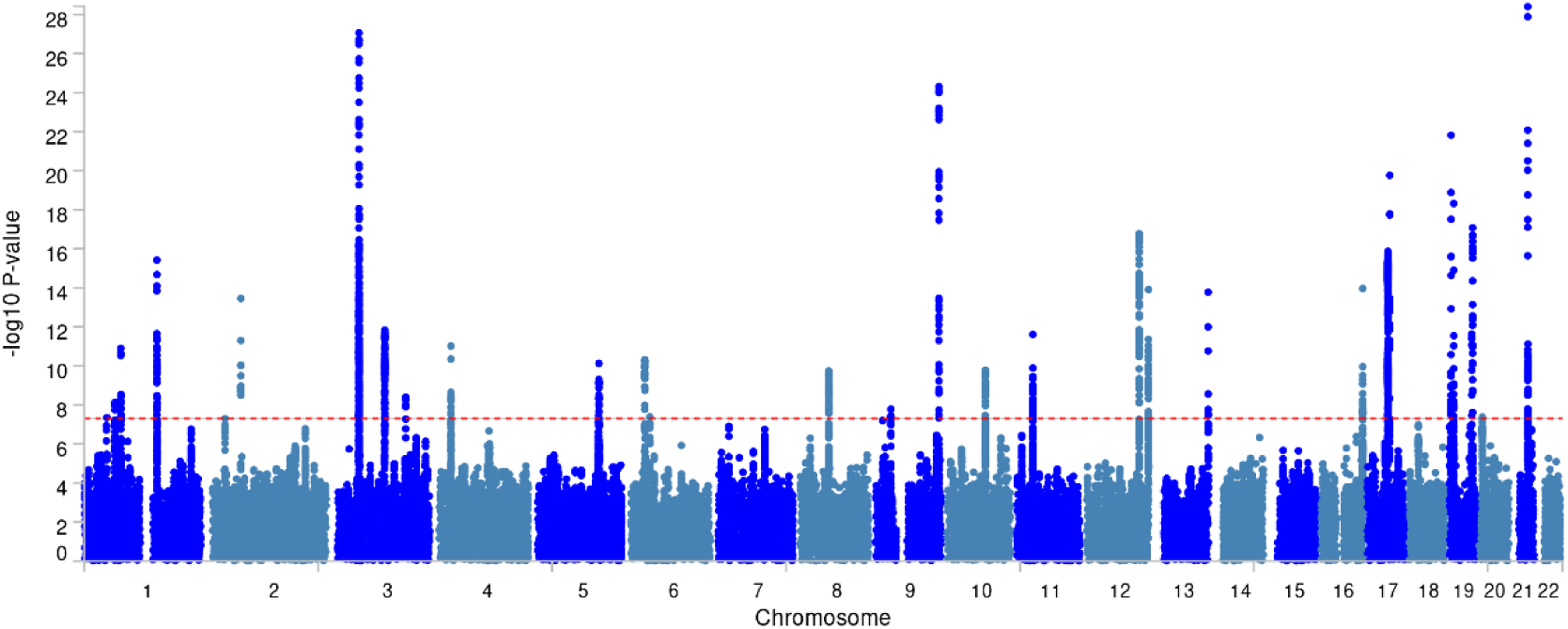
Manhattan Plots of the Differential GWAS. Manhattan plots depict P-values from the differential GWAS comparing hospitalized influenza and COVID-19.

**Table 4.**
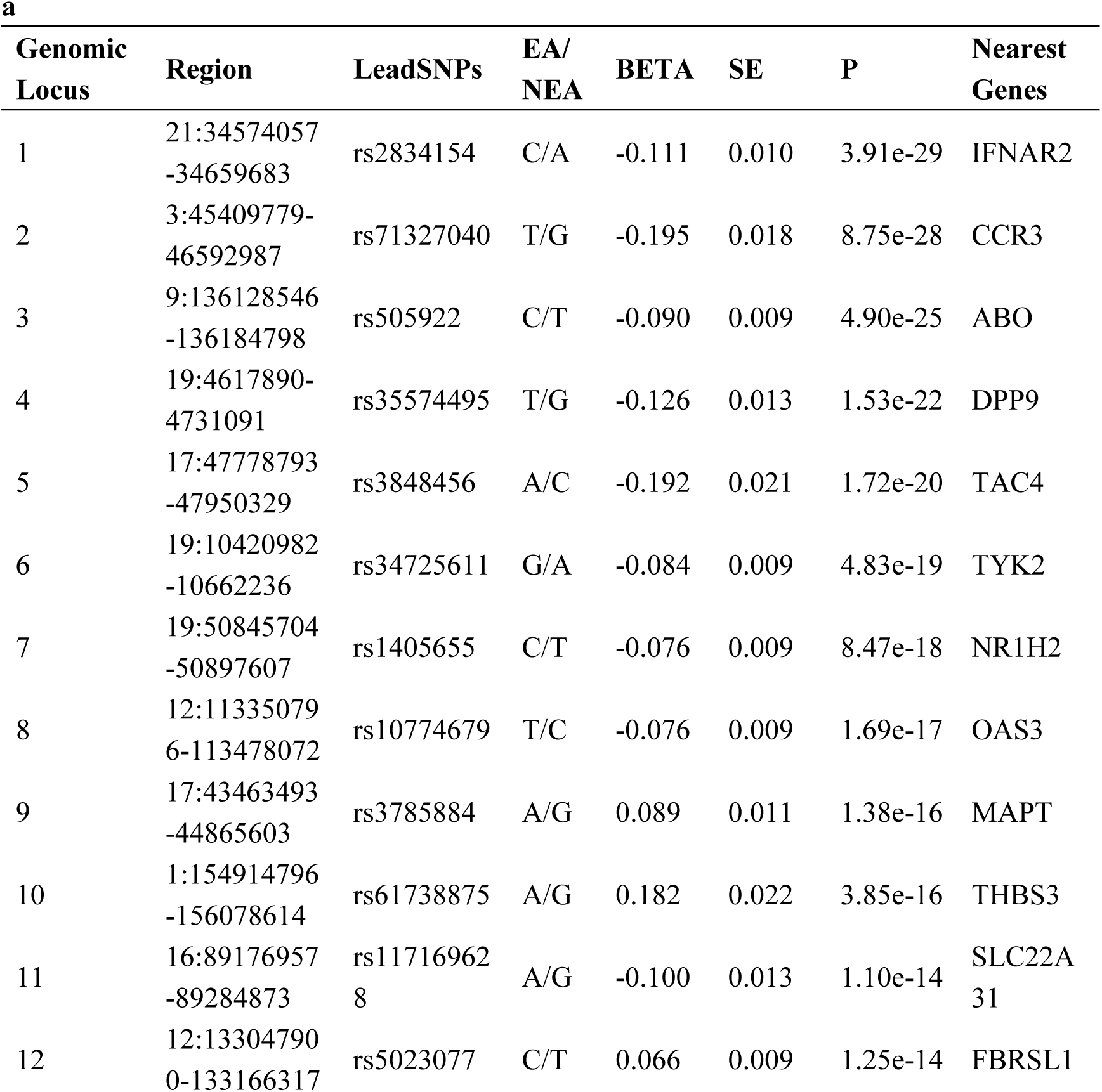
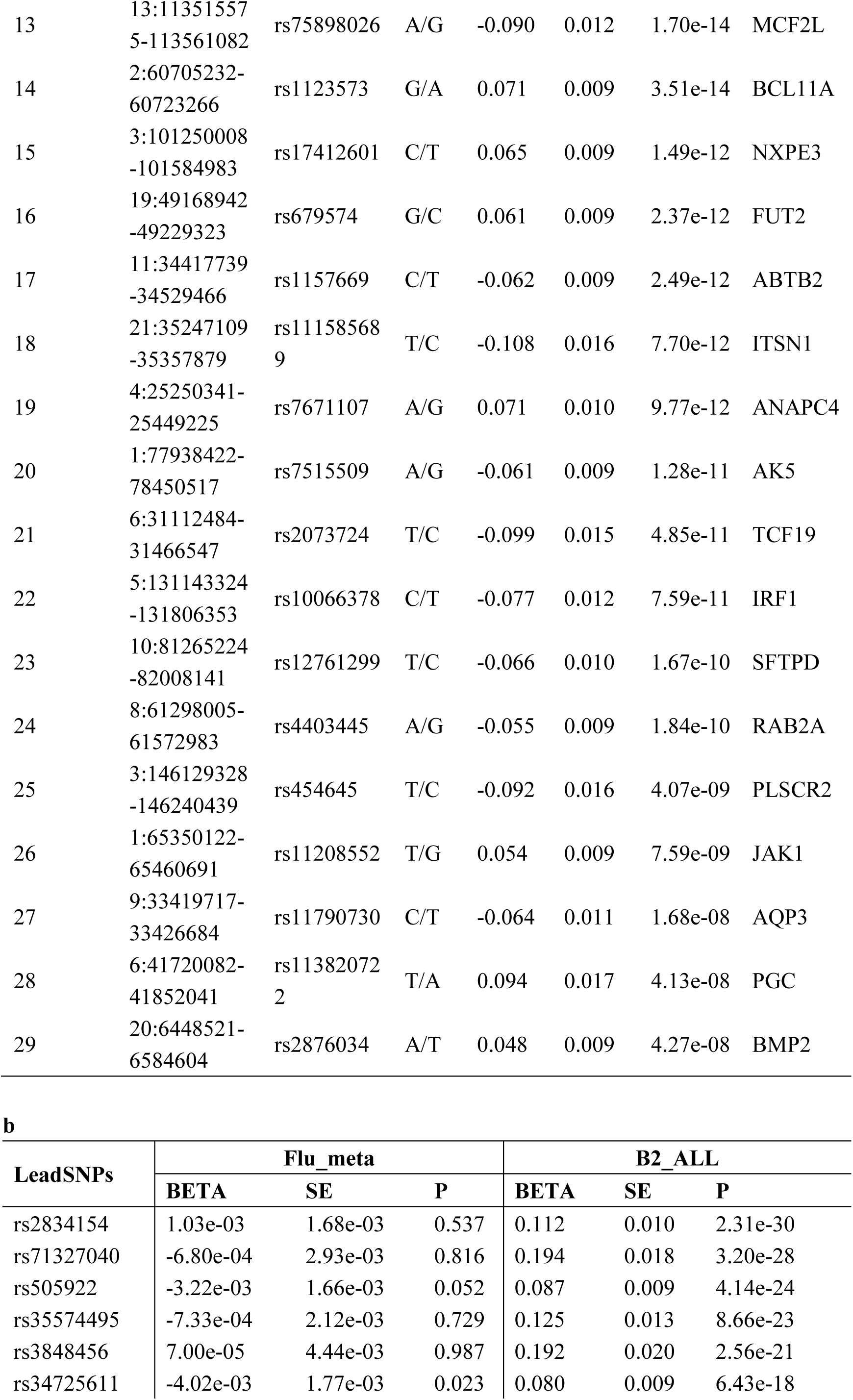
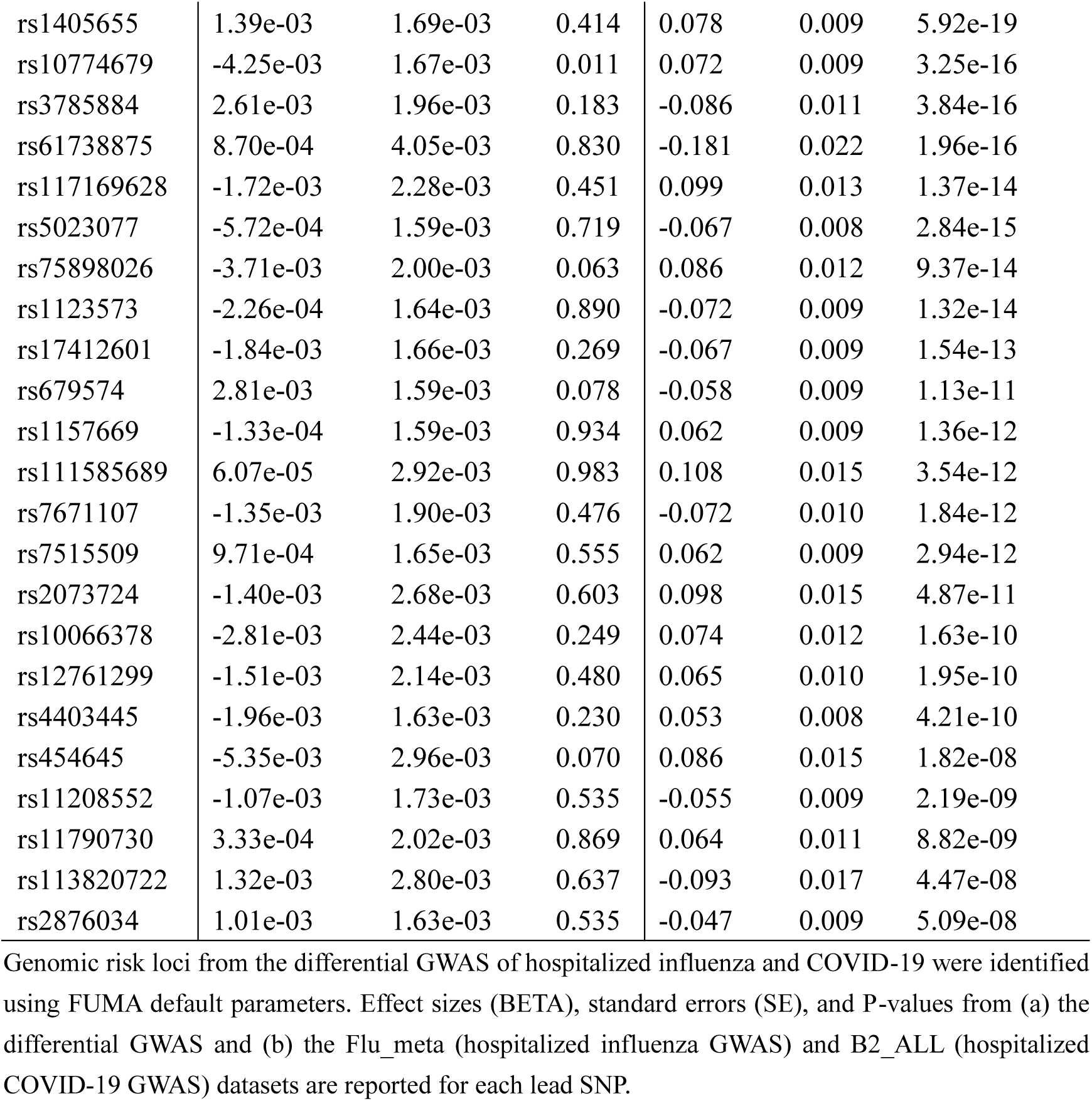
Genomic Risk Loci of the Differential GWAS a.

Additionally, we prioritized 29 genes located closest to each risk locus using the prioritization framework described in the Methods section (Table 5). Multiple analytical approaches—MAGMA, TWAS, spTWAS, and PWAS—were applied to the DDx-derived differential GWAS (see Supplementary Tables 6, 7, 8, and 9 for detailed results). Among the prioritized genes, RAB2A and FBRSL1 emerged as the most promising candidates, as they fulfilled the highest number of prioritization criteria, making them strong candidates for distinguishing between hospitalized influenza and COVID-19. The statistical results of gene-based tests for the top five genes with the highest prioritization scores are presented in Table 6. All identified genes were significantly associated exclusively with hospitalized COVID-19, with no significant associations found in hospitalized influenza. Similarly, three gene sets (Table 7) and two tissues (Figure 4) were significantly associated with hospitalized COVID-19 but not with influenza, as determined through MAGMA analysis.

**Figure 4.**
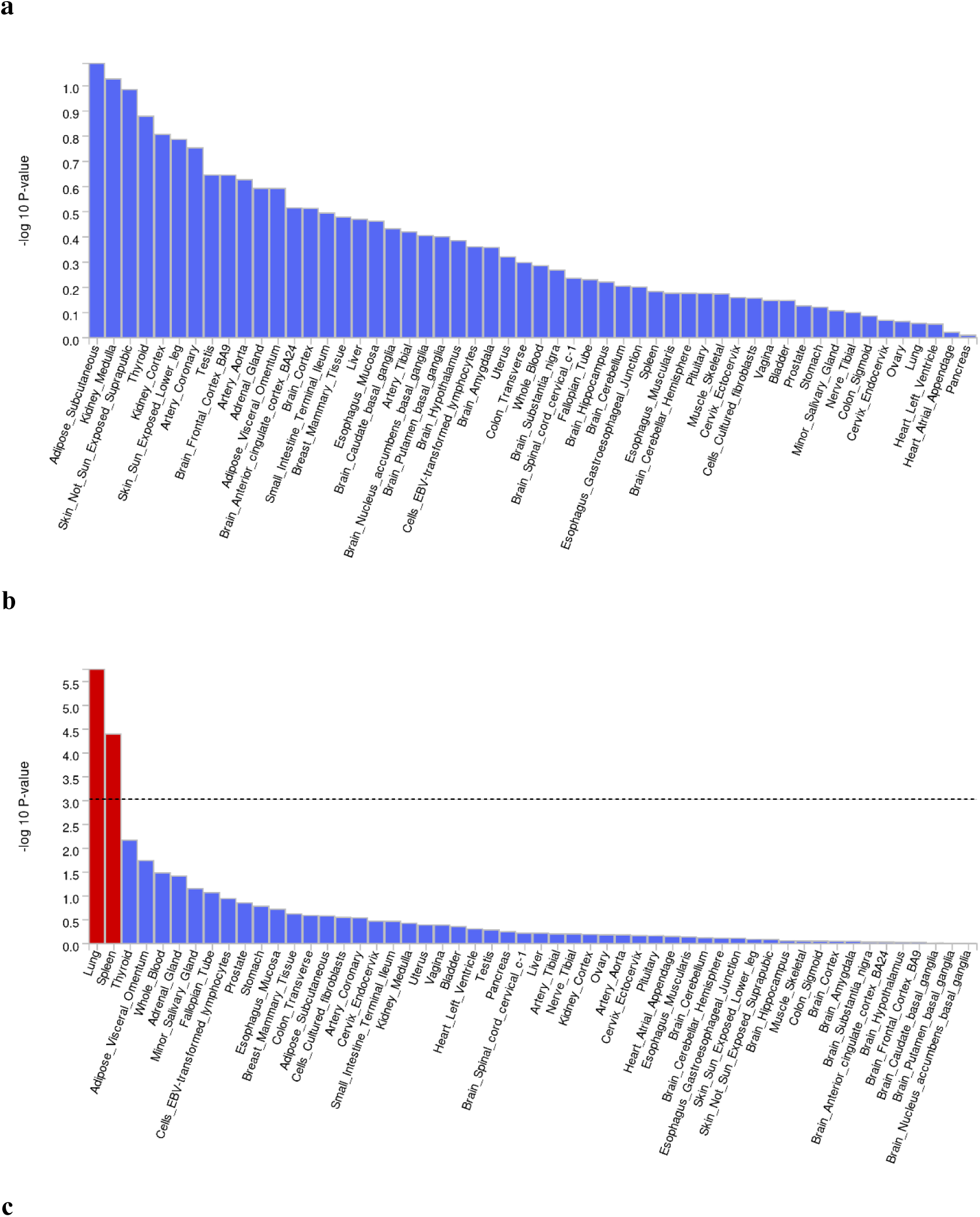
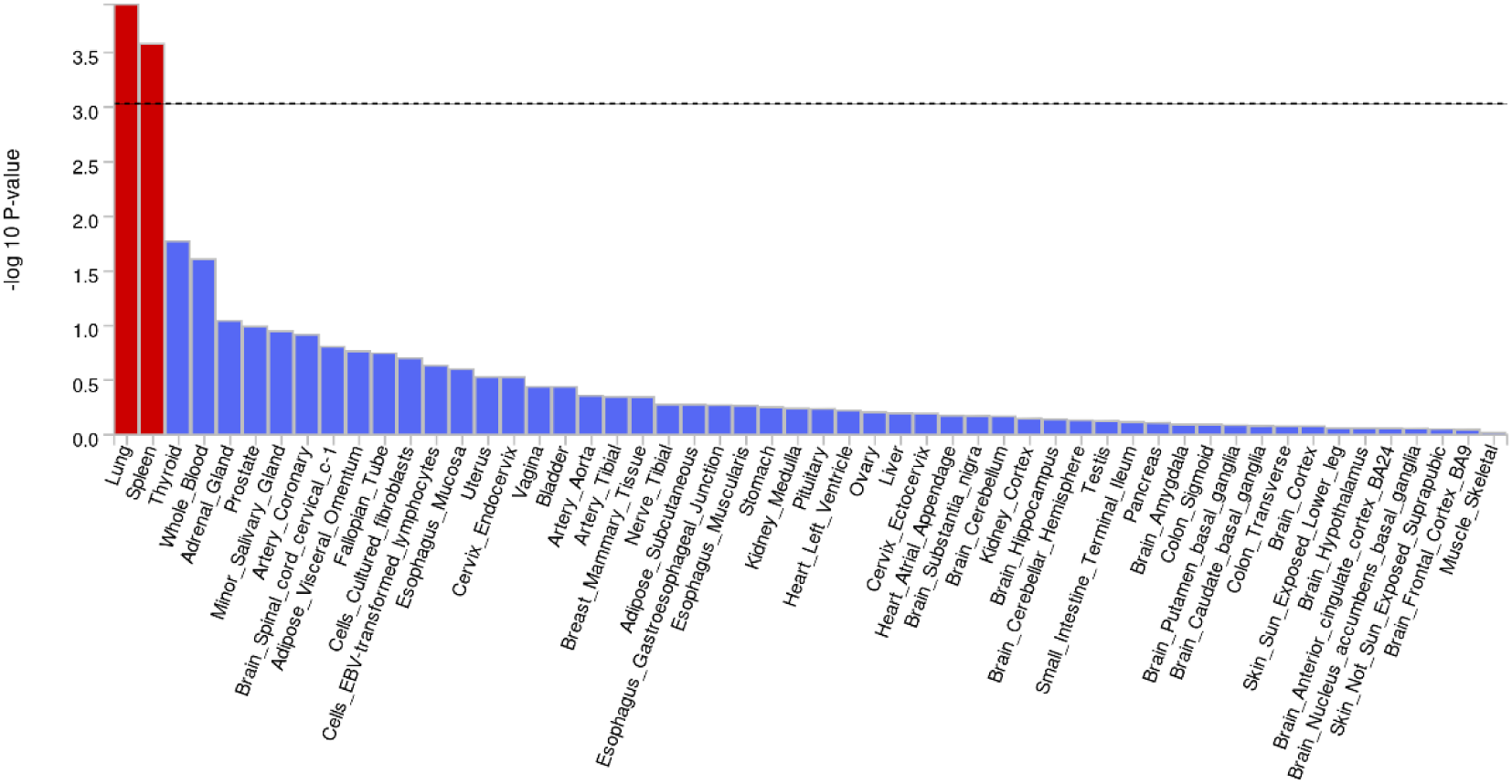
Tissues Differentially Associated with Hospitalized Influenza and COVID-19. Tissue enrichment analyses were conducted for (a) hospitalized influenza, (b) hospitalized COVID-19, and (c) the differential GWAS of hospitalized influenza and COVID-19 using MAGMA gene-property analysis in FUMA. Significant tissues are highlighted in red.

**Table 5.** Prioritization of Differential Genes.

| Gene | Open Targets |  | FUMA SNP2GENE |  |  |  | Gene-based Tests |  |  |  | PheWAS |  | Total |
| --- | --- | --- | --- | --- | --- | --- | --- | --- | --- | --- | --- | --- | --- |
|  | Closest Gene | Highest V2G | Strongest eQTL | Highest CADD | Lowest RegulomeDB | Nonsynonymous Variant | MAGMA | TWAS | spTWAS | PWAS | Immune | Respiratory |  |
| RAB2A | 1 | 1 | 1 | 1 | 1 | 0 | 1 | 1 | 1 | 1 | 1 | 0 | 10 |
| FBRSL1 | 1 | 1 | 1 | 1 | 1 | 1 | 1 | 1 | 1 | 0 | 1 | 0 | 10 |
| DPP9 | 1 | 1 | 1 | 1 | 1 | 0 | 1 | 1 | 1 | 0 | 0 | 1 | 9 |
| MAPT | 1 | 1 | 1 | 1 | 1 | 1 | 1 | 0 | 0 | 0 | 1 | 1 | 9 |
| TCF19 | 1 | 1 | 0 | 0 | 1 | 1 | 1 | 1 | 1 | 0 | 1 | 1 | 9 |
| IFNAR2 | 1 | 1 | 0 | 1 | 1 | 1 | 1 | 1 | 1 | 0 | 0 | 0 | 8 |
| ABO | 1 | 1 | 0 | 1 | 1 | 0 | 1 | 1 | 1 | 1 | 0 | 0 | 8 |
| TYK2 | 1 | 1 | 1 | 1 | 0 | 1 | 1 | 0 | 1 | 0 | 1 | 0 | 8 |
| THBS3 | 1 | 0 | 1 | 1 | 1 | 0 | 1 | 0 | 1 | 0 | 1 | 1 | 8 |
| SLC22A31 | 1 | 1 | 1 | 1 | 0 | 1 | 1 | 1 | 0 | 0 | 1 | 0 | 8 |
| BCL11A | 1 | 1 | 1 | 1 | 1 | 0 | 1 | 0 | 0 | 0 | 1 | 1 | 8 |
| AK5 | 1 | 1 | 1 | 1 | 1 | 0 | 1 | 0 | 0 | 0 | 1 | 1 | 8 |
| SFTPD | 1 | 0 | 1 | 1 | 0 | 1 | 1 | 0 | 1 | 0 | 1 | 1 | 8 |
| JAK1 | 1 | 1 | 1 | 1 | 1 | 0 | 1 | 1 | 0 | 0 | 1 | 0 | 8 |
| CCR3 | 1 | 1 | 1 | 0 | 1 | 0 | 1 | 1 | 0 | 0 | 1 | 0 | 7 |
| NR1H2 | 1 | 0 | 1 | 0 | 1 | 0 | 1 | 0 | 1 | 0 | 1 | 0 | 6 |
| NXPE3 | 1 | 1 | 1 | 0 | 1 | 0 | 1 | 0 | 0 | 0 | 1 | 0 | 6 |
| FUT2 | 1 | 0 | 1 | 1 | 0 | 0 | 1 | 0 | 0 | 0 | 1 | 1 | 6 |
| ITSN1 | 1 | 1 | 0 | 0 | 0 | 0 | 1 | 0 | 1 | 0 | 1 | 1 | 6 |
| TAC4 | 1 | 0 | 1 | 0 | 0 | 0 | 1 | 0 | 0 | 0 | 1 | 1 | 5 |
| OAS3 | 1 | 0 | 1 | 0 | 0 | 0 | 1 | 1 | 0 | 0 | 1 | 0 | 5 |
| MCF2L | 1 | 0 | 1 | 1 | 1 | 0 | 0 | 0 | 0 | 0 | 1 | 0 | 5 |
| ANAPC4 | 1 | 1 | 1 | 0 | 0 | 0 | 1 | 0 | 1 | 0 | 0 | 0 | 5 |
| IRF1 | 1 | 0 | 1 | 0 | 0 | 0 | 1 | 0 | 0 | 0 | 1 | 1 | 5 |
| BMP2 | 1 | 1 | 1 | 0 | 0 | 0 | 1 | 0 | 0 | 0 | 1 | 0 | 5 |
| ABTB2 | 1 | 0 | 1 | 0 | 0 | 0 | 0 | 1 | 0 | 0 | 0 | 1 | 4 |
| PLSCR2 | 1 | 0 | 1 | 0 | 0 | 0 | 0 | 1 | 0 | 0 | 0 | 1 | 4 |
| AQP3 | 1 | 0 | 1 | 0 | 0 | 0 | 0 | 1 | 0 | 0 | 1 | 0 | 4 |
| PGC | 1 | 0 | 1 | 0 | 0 | 0 | 0 | 1 | 0 | 0 | 1 | 0 | 4 |

Genes are scored against criteria identical to Table 3, with TWAS/spTWAS analyses restricted to lung and whole blood tissues.

**Table 6.**
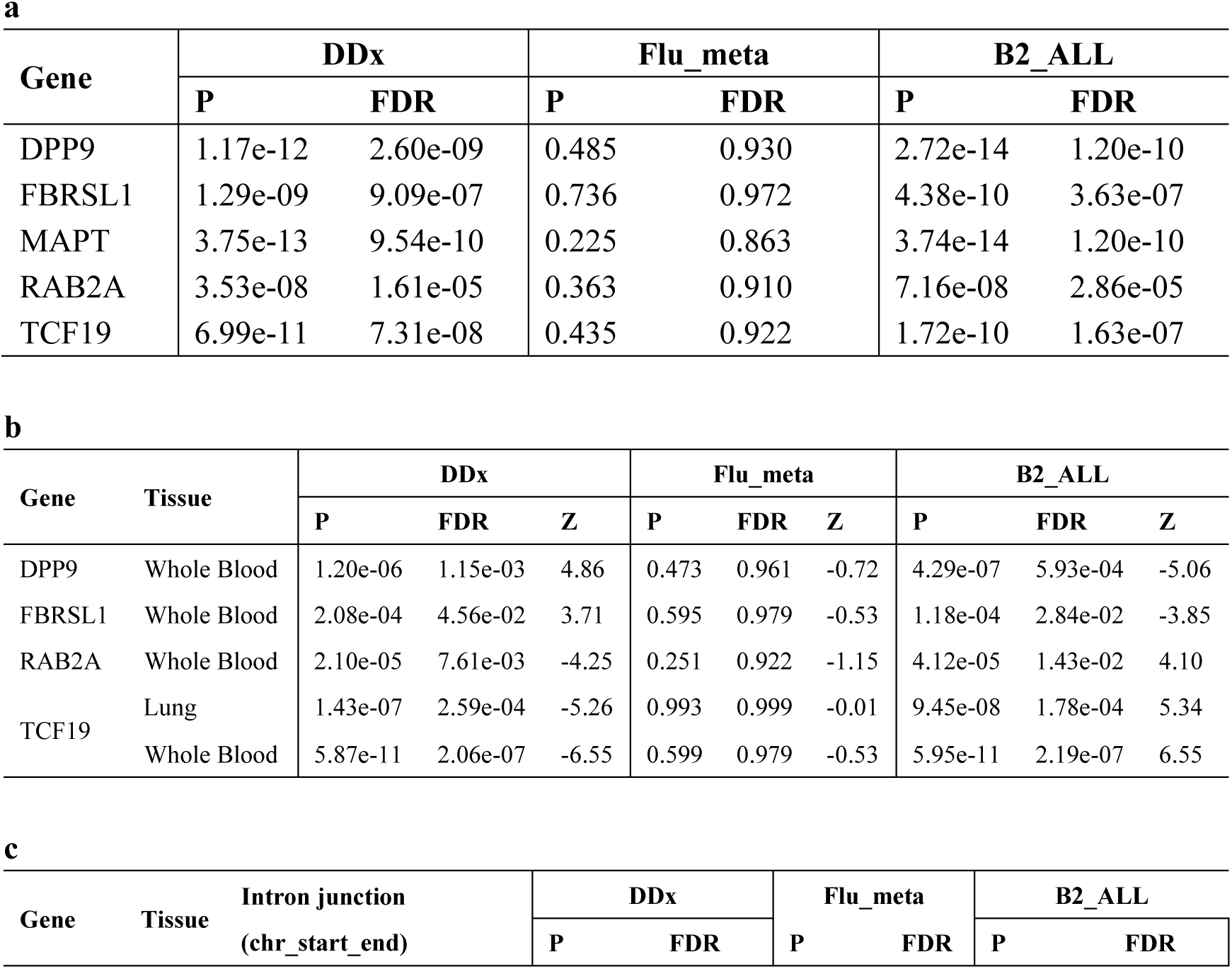
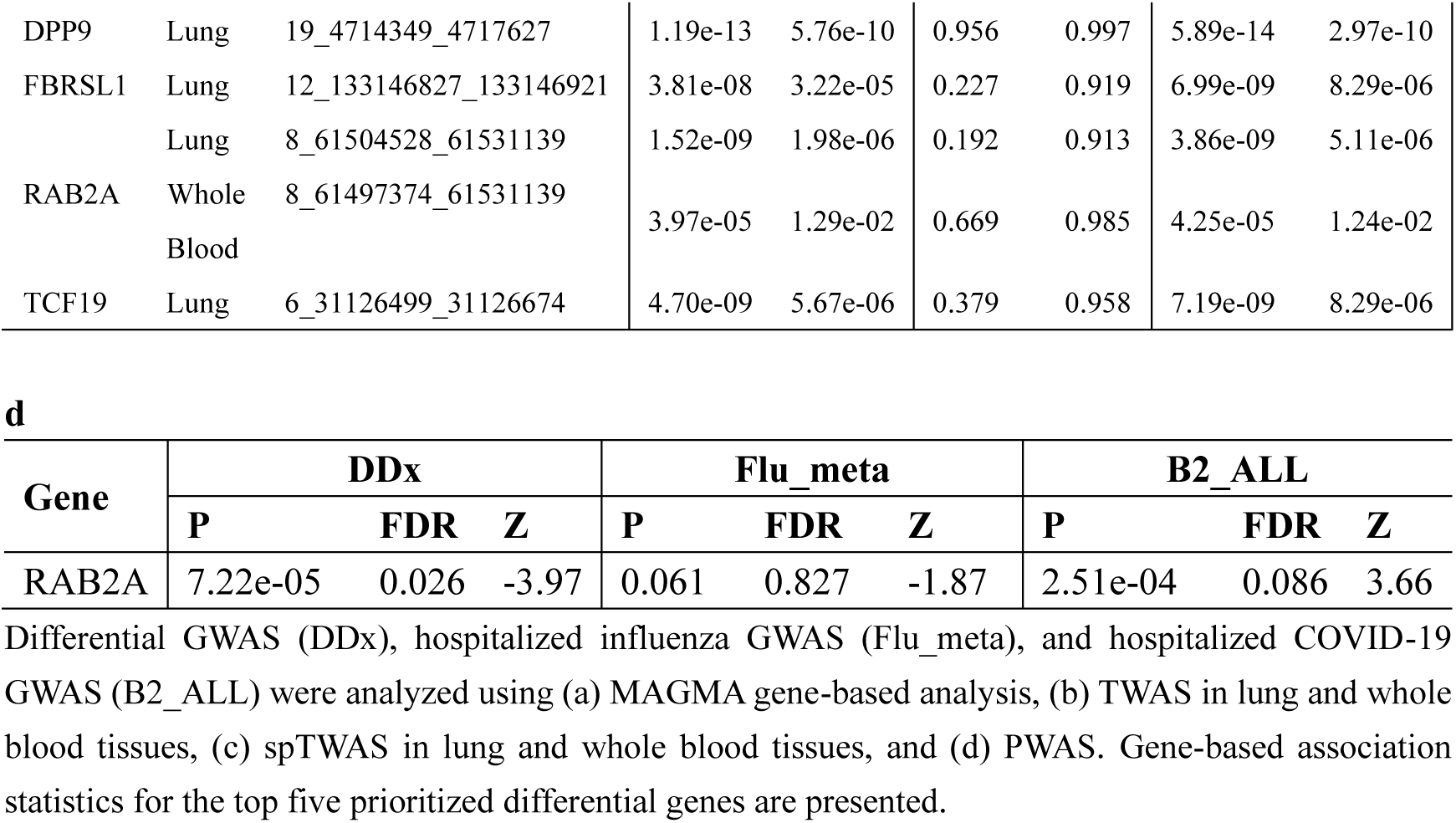
Gene-Based Test Statistics for the Top Five Prioritized Differential Genes a.

**Table 7.** Gene Sets Differentially Associated with Hospitalized Influenza and COVID-19.

| Gene Set | DDx |  | Flu_meta |  | B2_ALL |  |
| --- | --- | --- | --- | --- | --- | --- |
|  | P | FDR | P | FDR | P | FDR |
| Defective CSF2RB causes SMDP5 | 2.07e-10 | 3.52e-06 | 0.594 | 0.908 | 3.32e-09 | 2.82e-05 |
| Diseases associated with surfactant metabolism | 1.79e-09 | 3.04e-05 | 0.431 | 0.873 | 6.31e-08 | 2.68e-04 |
| Nikolsky breast cancer 1q21 amplicon | 1.09e-06 | 0.019 | 0.392 | 0.857 | 3.73e-08 | 2.11e-04 |

Differential GWAS (DDx), hospitalized influenza GWAS (Flu\_meta), and hospitalized COVID-19 GWAS (B2\_ALL) were analyzed using MAGMA gene-set analysis implemented in FUMA.

### Colocalization Analysis of Hospitalized Influenza and COVID-19 at the SNP and Gene Levels

To identify shared SNPs between hospitalized influenza and COVID-19, we applied the cofdr algorithm using Z-scores of overlapping variants from the respective GWAS as input. The top 10 SNPs with the highest posterior probability of association (PPA), as determined by cofdr, are listed in Table 8. However, none of these SNPs met the significance threshold of PPA ≥ 0.9. Notably, rs2278442-A exhibited the highest PPA, though at a moderate level of 0.564. This SNP was nominally significant in hospitalized influenza (P = 5.69×10⁻⁴) and suggestively significant in hospitalized COVID-19 (P = 1.58×10⁻⁶). It was associated with an increased risk of hospitalized influenza but had a protective effect against hospitalized COVID-19. The DDx algorithm also identified rs2278442-A as a significant differential SNP at a suggestive significance level (P = 1.03×10⁻⁷).

**Table 8.** Top 10 Colocalized SNPs in Hospitalized Influenza and COVID-19.

| SNP | EA/<br>NEA | Nearest<br>Genes | PPA | Flu_meta |  |  | B2_ALL |  |  |
| --- | --- | --- | --- | --- | --- | --- | --- | --- | --- |
|  |  |  |  | BETA | SE | P | BETA | SE | P |
| rs2278442 | A/G | ICAM3 | 0.564 | 0.006 | 0.002 | 5.69e-04 | -0.047 | 0.010 | 1.58e-06 |
| rs74364007 | T/C | OTOP1 | 0.501 | 0.016 | 0.004 | 1.58e-04 | -0.093 | 0.026 | 2.95e-04 |
| rs114486343 | A/G | TMEM128 | 0.456 | 0.018 | 0.004 | 6.08e-05 | -0.085 | 0.026 | 1.33e-03 |
| rs62329576 | A/G | SLC12A7 | 0.455 | 0.008 | 0.002 | 5.61e-05 | 0.050 | 0.017 | 3.58e-03 |
| rs9260118 | T/C | HLA-A | 0.445 | 0.006 | 0.002 | 2.58e-04 | -0.047 | 0.013 | 4.86e-04 |
| rs56395638 | G/A | ST6GAL1 | 0.423 | -0.012 | 0.002 | 7.33e-07 | 0.032 | 0.016 | 4.57e-02 |
| rs55658631 | A/G | ST6GAL1 | 0.410 | -0.012 | 0.002 | 7.33e-07 | 0.031 | 0.016 | 5.18e-02 |
| rs2735111 | A/G | HLA-A | 0.409 | 0.006 | 0.002 | 2.59e-04 | -0.045 | 0.013 | 9.00e-04 |
| rs9260119 | T/A | HLA-A | 0.408 | 0.006 | 0.002 | 2.59e-04 | -0.044 | 0.013 | 9.05e-04 |
| rs28718349 | G/A | STK11 | 0.405 | 0.006 | 0.002 | 9.66e-05 | -0.034 | 0.011 | 1.98e-03 |

Cofdr was applied to identify SNPs colocalized between the Flu\_meta (hospitalized influenza GWAS) and B2\_ALL (hospitalized COVID-19 GWAS) datasets. The table lists the top 10 SNPs with the highest posterior probability (PPA), along with their effect sizes (BETA), standard errors (SE), and P-values from the Flu\_meta and B2\_ALL GWAS.

For gene-level colocalization analysis, we used Z-scores derived from MAGMA, TWAS, spTWAS, and PWAS analyses based on GWAS data for hospitalized influenza and COVID-19. Applying a stringent significance threshold of PPA ≥ 0.9, we identified ST6GAL1 and ICAM5 as colocalized genes associated with both conditions through MAGMA-based cofdr and PWAS-based cofdr, respectively (Table 9a; Table 9d). Relaxing the PPA threshold yielded additional findings, identifying 32, 3, 22, and 5 shared genes through MAGMA-based cofdr, TWAS-based cofdr in lung tissue and EBV-transformed lymphocytes, spTWAS-based cofdr in lung and whole blood tissues, and PWAS-based cofdr, respectively (Table 9).

**Table 9.** Colocalized Genes in Hospitalized Influenza and COVID-19.

**a**
| Gene | PPA | Flu_meta |  | B2_ALL |  |
| --- | --- | --- | --- | --- | --- |
|  |  | P | FDR | P | FDR |
| ST6GAL1 | 0.971 | 1.14e-09 | 2.02e-05 | 3.60e-03 | 0.150 |
| **THBS3** | 0.877 | 6.08e-04 | 0.310 | 2.47e-07 | 8.22e-05 |
| NEIL3 | 0.857 | 3.60e-04 | 0.279 | 2.69e-03 | 0.130 |
| TCEA2 | 0.843 | 2.82e-04 | 0.279 | 1.64e-02 | 0.322 |
| BCL2L15 | 0.823 | 5.05e-05 | 0.133 | 7.09e-02 | 0.542 |
| PPP1R13B | 0.820 | 5.28e-04 | 0.304 | 6.46e-03 | 0.204 |
| KRTCAP2 | 0.816 | 1.10e-03 | 0.345 | 1.80e-07 | 6.34e-05 |
| PPP1CC | 0.800 | 4.91e-03 | 0.490 | 7.21e-04 | 0.062 |
| HIST1H1T | 0.799 | 7.46e-04 | 0.324 | 8.04e-03 | 0.228 |
| SSR2 | 0.778 | 1.01e-03 | 0.334 | 1.43e-02 | 0.301 |
| RSBN1 | 0.778 | 2.61e-04 | 0.279 | 4.52e-02 | 0.471 |
| PHTF1 | 0.772 | 2.67e-04 | 0.279 | 5.17e-02 | 0.492 |
| YY1AP1 | 0.767 | 3.87e-03 | 0.467 | 4.49e-04 | 0.047 |
| PTPN22 | 0.764 | 2.28e-04 | 0.279 | 6.45e-02 | 0.532 |
| DGKB | 0.759 | 3.53e-04 | 0.279 | 3.78e-02 | 0.441 |
| DUOXA2 | 0.749 | 2.24e-03 | 0.401 | 6.32e-03 | 0.202 |
| RAD23B | 0.742 | 2.03e-03 | 0.401 | 8.53e-03 | 0.236 |
| HIST1H2BJ | 0.733 | 4.44e-03 | 0.488 | 3.72e-03 | 0.153 |
| CYP19A1 | 0.733 | 8.77e-04 | 0.334 | 3.87e-02 | 0.446 |
| MTX1 | 0.730 | 2.10e-04 | 0.279 | 6.63e-12 | 8.84e-09 |
| MUC1 | 0.729 | 3.00e-03 | 0.431 | 2.71e-08 | 1.25e-05 |
| TRIM38 | 0.727 | 4.82e-05 | 0.133 | 9.88e-02 | 0.590 |
| CRNKL1 | 0.725 | 3.47e-04 | 0.279 | 8.81e-02 | 0.571 |
| AP4B1 | 0.723 | 5.21e-05 | 0.133 | 2.06e-01 | 0.703 |
| OXR1 | 0.722 | 2.39e-03 | 0.401 | 1.02e-02 | 0.255 |
| ICAM5 | 0.720 | 8.82e-03 | 0.593 | 8.94e-05 | 0.014 |
| LTF | 0.717 | 2.81e-02 | 0.713 | 2.63e-04 | 0.032 |
| ZFYVE21 | 0.714 | 5.45e-03 | 0.490 | 1.26e-03 | 0.087 |
| **ABTB2** | 0.712 | 4.92e-03 | 0.490 | 4.20e-03 | 0.166 |
| OAS1 | 0.705 | 1.05e-02 | 0.625 | 3.44e-13 | 6.86e-10 |
| EFNA1 | 0.703 | 1.84e-03 | 0.382 | 3.84e-06 | 9.58e-04 |
| MSANTD4 | 0.701 | 4.40e-04 | 0.304 | 9.05e-02 | 0.575 |

**b**
| Gene | PPA | Tissue | Flu_meta |  |  | B2_ALL |  |  |
| --- | --- | --- | --- | --- | --- | --- | --- | --- |
|  |  |  | P | FDR | Z | P | FDR | Z |
| MUC1 | 0.721 | EBV-transformed lymphocytes | 3.11e-04 | 0.289 | -3.61 | 5.07e-04 | 0.098 | 3.48 |
| ADAM15 | 0.715 | Lung | 2.65e-03 | 0.572 | 3.01 | 1.83e-04 | 0.050 | -3.74 |
| **OAS3** | 0.705 | EBV-transformed lymphocytes | 9.72e-03 | 0.624 | -2.59 | 2.24e-13 | 1.18e-09 | 7.33 |

**c**
| Gene | PPA | Tissue | Intron junction<br>(chr_start_end) | Flu_meta |  | B2_ALL |  |
| --- | --- | --- | --- | --- | --- | --- | --- |
|  |  |  |  | P | FDR | P | FDR |
| TCEA2 | 0.851 | Lung | 20_62694737_62695245 | 5.33e-04 | 0.488 | 3.11e-03 | 0.172 |
| **THBS3** | 0.850 | Lung | 1_155176197_155177589 | 3.03e-04 | 0.446 | 4.68e-03 | 0.216 |
| PLEKHA4 | 0.813 | Lung | 19_49364771_49364877 | 5.33e-03 | 0.747 | 2.52e-05 | 0.006 |
| MUC1 | 0.800 | Lung | 1_155162038_155162577 | 3.11e-04 | 0.446 | 9.45e-04 | 0.095 |
| ATP11A | 0.783 | Lung | 13_113534963_113536190 | 3.00e-02 | 0.798 | 1.02e-11 | 0.000 |
| OAS1 | 0.773 | Lung | 12_113355505_113356402 | 1.24e-02 | 0.766 | 8.32e-14 | 0.000 |
| FHL3 | 0.773 | Lung | 1_38464820_38464929 | 1.96e-03 | 0.669 | 3.08e-03 | 0.172 |
| TMEM50B | 0.770 | Lung | 21_34828404_34832720 | 5.67e-03 | 0.747 | 5.91e-04 | 0.069 |
| METTL8 | 0.757 | Lung | 2_172188377_172193963 | 2.53e-03 | 0.702 | 3.01e-03 | 0.170 |
| **THBS3** | 0.756 | Whole Blood | 1_155176197_155177589 | 3.03e-04 | 0.520 | 4.68e-03 | 0.233 |
| SLC12A8 | 0.733 | Lung | 3_124829179_124837613 | 2.68e-03 | 0.714 | 4.64e-03 | 0.216 |
| **ANAPC4** | 0.729 | Lung | 4_25394043_25395427 | 1.37e-02 | 0.766 | 1.18e-04 | 0.021 |
| PPP1R13B | 0.724 | Lung | 14_104216271_104219337 | 4.23e-04 | 0.465 | 2.75e-02 | 0.466 |
| FDX2 | 0.717 | Lung | 19_10426168_10426381 | 2.54e-02 | 0.790 | 7.56e-11 | 0.000 |
| ZCCHC4 | 0.715 | Lung | 4_25366788_25370651 | 1.47e-02 | 0.766 | 8.42e-05 | 0.016 |
| PNPO | 0.708 | Lung | 17_46022081_46022925 | 2.16e-02 | 0.790 | 1.97e-05 | 0.005 |
| NCOR1 | 0.705 | Lung | 17_16052831_16054904 | 2.36e-02 | 0.790 | 8.11e-06 | 0.003 |

**d**
| Gene | PPA | Flu_meta |  |  | B2_ALL |  |  |
| --- | --- | --- | --- | --- | --- | --- | --- |
|  |  | P | FDR | Z | P | FDR | Z |
| ICAM5 | 0.955 | 6.13e-04 | 0.275 | 3.43 | 8.63e-04 | 0.103 | -3.33 |
| GSTM1 | 0.844 | 2.28e-03 | 0.419 | 3.05 | 3.80e-03 | 0.217 | 2.89 |
| STAT3 | 0.812 | 8.29e-03 | 0.539 | 2.64 | 1.98e-03 | 0.152 | -3.09 |
| CTSH | 0.793 | 1.50e-02 | 0.564 | 2.43 | 2.93e-04 | 0.086 | -3.62 |
| NPNT | 0.777 | 2.22e-02 | 0.615 | 2.29 | 5.44e-06 | 0.007 | 4.55 |

## Discussion

We conducted a binary genome-wide association study (GWAS) and a time-to-event GWAS to investigate the genetic factors influencing hospitalized influenza, utilizing data from the UK Biobank and FinnGen. Our analyses identified four genomic risk loci, with the strongest association observed at the ST6GAL1 gene. Further differentiation analysis comparing hospitalized influenza and COVID-19 GWAS findings revealed distinct genetic architectures, with 29 differentially associated loci and genes, as well as three gene sets and two tissue types specific to COVID-19. Additionally, colocalization analysis suggested shared genetic mechanisms between these respiratory infections, highlighting ST6GAL1 and ICAM5 as potential key candidates.

### ST6GAL1 as a Key Locus in Influenza Pathogenesis

ST6GAL1 consistently emerged as a significant locus in both the binary and meta-analyzed time-to-event GWAS, making it the most promising candidate associated with hospitalized influenza. The lead SNP within this locus, rs13088725, was particularly notable for its strong association with a reduced risk of hospitalization and the lowest p-value among all examined SNPs, showing its potential role in genetic susceptibility to influenza. Moreover, rs13088725 is associated with mean platelet volume, an inflammatory marker, suggesting its involvement in immune response and inflammation. This variant is also significantly linked to lower ST6GAL1 expression in blood tissue, offering insights into a possible genetic mechanism influencing influenza risk. Functionally, ST6GAL1 regulates sialic acid receptor synthesis, which serves as a binding target for influenza viruses, facilitating viral entry into host cells^44, 45^. Suppressing ST6GAL1 expression or activity has been shown to reduce viral entry and replication in epithelial cells^46–48^ and infant mice^48^, indicating its potential as a therapeutic target.

### Roles of SPATS2L and AASDHPPT in Immune and Metabolic Pathways

SPATS2L ranked second in our integrative prioritization framework. It is upregulated in response to type I interferons (IFN) and plays a role in autoimmunity^39, 40^. The IFN signaling pathway is crucial for both innate and adaptive immune responses against viral infections, including influenza, by triggering antiviral protein production^49^. Additionally, SPATS2L was identified as a key biomarker with high fold-change in gene expression during influenza challenge tests^50^, suggesting its potential involvement in disease pathophysiology. Its differential expression in the whole blood of pediatric influenza patients^51^ further supports its role in antiviral immune responses. Moreover, its identification as a causal gene for lung function^43^ and bronchodilator response in asthma^52^ highlights its broader impact on respiratory health. While direct experimental evidence linking SPATS2L to influenza remains limited, its role in IFN signaling and respiratory function suggests intriguing diagnostic and therapeutic possibilities.

Another significant locus identified in the binary GWAS was rs17093811, located near the AASDHPPT gene. This gene encodes a phosphopantetheinyl transferase enzyme involved in the biosynthesis of coenzyme A (CoA)^53^, a key factor in amino acid and lipid metabolism, which influences cellular energy production and utilization^54^. AASDHPPT also plays a role in mitochondrial fatty acid synthesis (mtFAS)^55–57^, an essential process for mitochondrial respiration and oxidative metabolism. Given its fundamental role in biosynthetic pathways, disruptions in AASDHPPT function may affect metabolic balance and indirectly influence susceptibility to influenza.

### UGT2B4 and Temporal Dynamics of Influenza Hospitalization

In the time-to-event GWAS, we identified a novel locus, rs6821234, near the UGT2B4 gene, with UGT2A1 and UGT2A2 as the closest neighboring genes. This locus is responsible for regulating isoform expression in the UGT2A and UGT2B gene families, which encode UDP-glucuronosyltransferase (UGT) enzymes essential for metabolizing various substances^58, 59^. The involvement of UGT2B4 in xenobiotic metabolism suggests that genetic variation in detoxification pathways may influence the timing or progression of severe influenza. The role of UGT2B4 in metabolizing estrogens and fatty acids^60^ raises intriguing questions about sex-specific or age-related differences in disease severity, requiring further investigation.

### Differentiated Genetic Architectures Between Influenza and COVID-19

In the differential GWAS, we identified 29 loci with lead SNPs reaching genome-wide significance in COVID-19 (excluding rs2876034 and rs12567716, which had P < 5 × 10⁻⁷) but not in hospitalized influenza, indicating the presence of genetic factors unique to COVID-19. Notably, RAB2A and FBRSL1 emerged as key differentiating genes, suggesting distinct roles in COVID-19 pathophysiology and potential therapeutic targets. RAB2A has a pro-viral role in COVID-19 by facilitating SARS-CoV-2 replication through interactions with viral proteins NSP7 and ORF3a^61^. Although FBRSL1 has not been previously associated with lung traits, phenome-wide association studies (PheWAS) linked it to red blood cells^62^, which are vital for oxygen transport, and to comorbidities commonly observed in COVID-19 patients, such as type 2 diabetes^63^ and cardiovascular disease^64, 65^. Aggregating gene-level differential effects further revealed three gene sets and two tissues (lung and spleen) uniquely associated with COVID-19, suggesting that genetic factors specific to COVID-19 may particularly influence immune and pulmonary functions.

### Shared Mechanisms and Therapeutic Implications

In the colocalization analysis, the top-ranking variant rs2278442-A exhibited opposing effects between hospitalized influenza and COVID-19 and was identified as a suggestively significant differential SNP by the DDx algorithm. PheWAS linked this variant to lower lymphocyte counts (P = 4 × 10⁻³²)^66^, an indicator of weakened immune response. Additionally, rs2278442-A is an intronic variant of the ICAM3 gene, which encodes intercellular adhesion molecule 3, a protein highly expressed in lymphocytes^35^ and essential for immune function. Given that rs2278442-A is an expression quantitative trait locus (eQTL) associated with higher ICAM3 expression^36^, this variant may influence immune response modulation and, consequently, differential susceptibility to influenza and COVID-19.

Gene-level colocalization analysis identified ST6GAL1 with the highest posterior probability of association (PPA) using MAGMA-based cofdr, aligning with its known role in viral pathogenesis and its upregulation in severe COVID-19 cases^67^. Additionally, ICAM5 was identified through PWAS-based cofdr and exhibited opposing associations between hospitalized influenza and COVID-19. Specifically, ICAM5 was protective against COVID-19, in line with previous Mendelian randomization studies^68–70^. Although its role in influenza is less understood, the involvement of other intercellular adhesion molecules in viral attachment and entry is well-documented^71^. For example, ICAM1 protein levels were upregulated following influenza virus infection and played a key role in immune defense^72^. Given their structural similarities^73^, we hypothesize that ICAM5 may also impact influenza severity through its role in viral infection.

Furthermore, among the 29 differential genes identified in the differential GWAS, four genes—THBS3, ABTB2, OAS3, and ANAPC4—were also detected in colocalization analysis with a more relaxed PPA threshold. These genes not only exhibit colocalization in both traits but also show differential associations, suggesting that they may modulate disease severity in opposing directions.

### Limitations and Future Directions

This study has some limitations. First, the meta-analyzed GWAS of hospitalized influenza focused only on White British and Finnish populations, limiting its generalizability. The statistical power was constrained by the relatively small sample size. Additionally, the reliance on ICD-10 codes for case definitions without distinguishing influenza subtypes introduces potential misclassification. Our analyses did not account for environmental or lifestyle factors, such as temperature, BMI, and smoking, which could influence influenza susceptibility. Another limitation is the reliance on bioinformatics tools without experimental validation, making it necessary to interpret differential and colocalized results with caution, particularly given the risks of pleiotropy and linkage disequilibrium.

## Conclusion

In conclusion, our binary and time-to-event GWAS analyses identified four genomic risk loci associated with hospitalized influenza. The strongest association with ST6GAL1 underscores its critical role in influenza pathogenesis and its potential as a therapeutic target. The prioritization of SPATS2L, AASDHPPT, and UGT2B4 suggests their involvement in influenza susceptibility. Differential analysis with hospitalized COVID-19 revealed distinct genetic architectures, with 29 differentially associated loci and genes. Colocalization analysis further identified shared genetic mechanisms, with ST6GAL1 and ICAM5 as key candidates. These findings contribute to a deeper understanding of the genetic basis of hospitalized influenza and its relationship with COVID-19, providing a foundation for future personalized treatment strategies.

## Data Availability

All data produced in the present study are available upon reasonable request to the authors.

## Notes

### Competing Interest Statement

The authors have declared no competing interest.

### Funding Statement

This study did not receive any funding.

### Author Declarations

The study used ONLY openly available human data that were originally located at UKBB, FinnGen and COVID HGI.

