## Supplementary material for "Genome-wide association studies of hospitalized influenza identify 4 risk loci and a relationship with COVID-19": Supp_Influenza_UKBB_GWAS.pdf

**SNP-based heritability.** To determine how important genetics is to the hospitalized influenza phenotype, we first estimated the SNP-based heritability (h2) on the liability scale using LDSC (Supplementary Table 1). There were not enough polygenic signals in the *Flu_ukbb* GWAS to get a reliable h2 estimate, as indicated by the mean chi-square of 1.003 (<1.02 suggested by LDSC). This could be attributed to the limited effective sample size in the cohort (Neff=4137). For the FinnGen data, h2 was 0.0452 (SE=0.0102), significantly different from 0 (P=9.36e-6). Meanwhile, a lower but more precise estimate of the SNP heritability was obtained in the meta-analysis (h2=0.0369, SE=0.0076) with the increasing sample size.

**Supplementary Table 1. Liability scale SNP-based heritability for hospitalized influenza**

| | **h2** | **h2.SE** | **h2. Z** | **h2. P** |
|---|---|---|---|---|
| UKBB | 0.0282 | 0.0464 | 0.608 | 0.543 |
| FinnGen | 0.0452 | 0.0102 | 4.431 | 9.36E-06 |
| Meta | 0.0369 | 0.0076 | 4.855 | 1.20E-06 |

"h2" means the liability scale heritability estimated by LDSC using hospitalized influenza GWAS from UKBB, FinnGen release 11, or meta-analysis. The corresponding standard error "h2.SE", z-score "h2.Z", and two-tailed p-value "h2.P" were also listed in the table.
